# Surveillance of the Impact of Antimicrobial Resistant infections in Immunosuppressed Children’s therapy: A systematic review

**DOI:** 10.1101/2023.04.14.23288454

**Authors:** Danielle Domo, Ivo Ngundu Woogeng

## Abstract

**Background:** Antimicrobial resistance (AMR) is a global problem. Resistant bacteria, for many reasons, do not cease to emerge and re-emerge. The impact of AMR on patient therapy is not scares in literature to date, but there is still much to do, mainly in pediatric settings. It is vital to assess the necessity of observing the impact of such infections on the clinical care trends to which some kids may already be subject, strengthening, improving, and, where necessary, implementing new age policies and regulations that may help contain the spread of AMR.

**Method:** We performed a yearlong review of literature on antimicrobial resistance in paediatrics immunosuppressed patients until September 2022. We draw-up a protocol of the review, to which we adhered rigorously, following the prescribed including and excluding criteria.

**Results:** From the 110 articles finally selected following the PRISMA workflow diagram, from which 29% of them were in majority randomized controlled trials studies, the remaining selection ranged from case controls to cohort studies, systematic reviews, controlled before and after reports trials, matched case-control and placebo reports trials and few not clearly reported research article types (15%).

**Conclusion:** The process yielded to the confirmation that there are literally good evidences of the serious lethal impact of resistant microbiological infections among immunocompromised children in and out of hospitals’ settings. What lacks the most are practical evidences of such damages both to the patient and to the public health sector, which can be conquer though, through well-programmed cohort–based studies.

## 1. Introduction

### 1.1. Rationale

According to the World Health Organisation’s (WHO), annual reports on microbial resistance, antimicrobial resistance (AMR) is a global threat (1–5). The European Committee on Antimicrobial Susceptibility Testing (EUCAST) defines AMR in two ways: firstly, by the population of bacteria that exist before exposure to the antimicrobial agent, and secondly, by adverse clinical outcomes related to uncontrolled infection, if a patient receives that antimicrobial (6).

A microorganism is resistant if it has an acquired or a genetic mutational type of reluctance mechanism to the drug in question (7). Clinical resistance, on the other hand, is defined by the level of antimicrobial activity associated with a high likelihood of therapeutic failure (8). Pathogens like Pseudomonas Pseudomonas Burkholderia sp. (15–17), Shigella sp. (18) and other panresistant bacteria (19), Pandorea sp. (20,21), among others gram-negative microorganisms (22–26), Staphylococci (MRSA (27), coagulase coagulase-negative Enterobacter sp.(29) and variants of the strep groups (30), have been identified so far as highly dangerous among immunocompromised patients, and also immunocompromised pediatric patients (31–33). Risk factors associated with bacterial resistance are diverse and often interconnected (34).

Exposure of a patient to certain drugs (35–37), and therapies (22,35), is likely to generate severe adverse consequences to infectious pathogens (38), just like in the case of mix infections (transfer of resistance mechanisms via bacterial phage), rending certain pathogens resistant to drugs they used to respond positively to (9,39–41). AMR develops when microorganisms (bacteria, fungi, viruses, or parasites) no longer respond to a drug it was previously sensitive to (3,42,43). This means that standard treatments no longer work; infections are more complicated and impossible to control; the risk of the spread of disease to others is higher; illness and hospital stays are prolonged, with added economic and social costs(44–46). The risk of death is increased in many cases, twice that of patients with infections caused by non-resistant bacteria (5,47). The clinical breakpoint seems to be that which explains or qualifies the microorganism’s change of resistance patterns throughout the years (39). The exact range of antibiotics use in pediatrics is still difficult to estimate(48). However, they are essential in treating most infections, meaning there is little to no choice in using or not using this treatment whenever necessary (49). The actual worry lies in the necessity to slow the ongoing dissemination of antimicrobial resistance and reduce the widespread impact on the subject’s health status today and in the near future (50), (51).

The current opinion is that not addressing AMR’s current and future problems with emphasis will result in severe health degradation (52–55). This is because most currently commercialized antibiotics, used for reverting the ongoing trends of resistances, are the third-level ones (56). Furthermore, these are often administrated only in hospital settings(35,57,58). As a result, they might be found less effective than the primary ones, more expensive, and, unfortunately, often associated with severe side effects on the patient’s status or therapy (4).

Reports indicate that AMR disproportionately affects high, low-income countries(44,59,60) with a relatively higher infection rate and limited access to effective therapies and funds for appropriate bacterial investigation and reporting from one place to the other (61). Therefore, creating protocols or re-enforce the existing ones overall or in specific settings, in particular, is highly necessary (61–64). The problem is so severe that it threatens the achievements of modern medicine, as a post-antibiotic era, in which common microbial infections and minor injuries will eventually cause death, is a genuine possibility. Therefore, a clinical surveillance system is necessary and should be monitored constantly. The main challenges in tackling antimicrobial resistance are social, ecological, intellectual, economic, and political (65). Unfortunately, middle-income countries hardly follow up with surveillance programs(51,66), are non-achieving good reporting (2), nor keep up with standard recommendations, such as reporting AMR with the type of sample of origin of the isolate, with accuracy (44).

In the routine microbiology laboratory, checking for antimicrobial resistance of bacteria to most antibiotics is a standard operating procedure (67,68). However, testing for antimicrobial resistance for most microbiologists has always been challenging and non-enthusiastic, especially when pediatric samples are involved. Though resistant bacteria have become an ordinary matter in clinical settings (1), they can still be controlled to the best of practice and be prevented by good clinical exercise and optimal turn-around time (69–73).

Yet, more must be done to provide public health services and hospitals with necessary information on the health, economic and social risks of dealing with resistant bacterial infections in any population type, among children and immune-suppressed children (74).

Therefore, the importance of this systematic review is to raise awareness of this latent problem before it becomes even more severe and help implement and improve antimicrobial stewardship programs worldwide.

### 1.2. Objectives

To provide a synthesis of what has been said in peer-reviewed literature, to date, on the impact of antibacterial resistance infections on immunosuppressed children. Provide necessary information on the patient’s mortality and morbidity after bacteremia, therapy, and final output of the combined care condition. The study will also be interested in the patient’s length of stay in the hospital after bacterial identification, therapy, and final output.

## 2. Methods

### 2.1. Eligibility criteria

The study selected as part of this systematic review followed the PECO based selection criteria:

­ Population: Humans, with emphasis on children aged 0 to 17 years old; immunosuppressed (suffering from genetic diseases causing immune depression, organ transplant patients, acquired immune suppressive disorders of any type, in and out of hospital patients)
­ Exposure: To antimicrobial-resistant infections
­ Confounders: Not applicable
­ Outcome: Mortality and hospital’s Length of stay (LoS) - Outcomes of treatment, either the resistant infections only on patient’s health status or effects of combined therapies.

All relevant reports will be included unless there is clear evidence of an update of an older edition. This study will give preference to peer-reviewed articles.

Only published manuscripts, conference abstracts, and outdated guidelines will be included.

Studies will be excluded because of outcomes that are not in line with the current study objectives (“wrong outcome”) and if the study of the impact of resistant infection on therapy is focused only on adult populations or cases.

### 2.2. Information sources

A first attempt to search for literature for the current study was on May 9^th,^ 2022, on the following database: PubMed, Embase, and Cochrane, Ovid’s “Bambino Gesù” Pediatric Hospital’s library platform in Rome. Secondarily to these conventional search strategies, a comprehensive grey literature search included Google search engine and websites from the ministries of health, healthcare services of Italy and Cameroon, institutes of public health, European Centre for Disease Prevention and Control (ECDC), World Health Organization (WHO), scientific societies in the field (including the European Society of Clinical Microbiology and Infectious Diseases (ESCMID), the International Society for Infectious Diseases (ISID), the International Epidemiological Association (IEA), the European Society of Intensive Care Medicine (ESICM) and the European Respiratory Society (ERS)). The search strategy used the following terms in English and local languages: ‘Antimicrobial resistance’ AND/OR ‘Hospital-associated’ OR ‘Hospital-acquired’ OR ‘Nosocomial’ AND ‘Surveillance’ AND ‘epidemiology’ OR ‘prevalence’ OR ‘incidence.’ In addition, the European Committee on Infection Control websites (EUCIC) of ESCM was consulted as an additional source to help us find specific publicly available documents that might have been missed out with our search strategy. However, only data from publicly available sources were included in the review.

### 2.3. Search strategy

This systematic review follows the PRISMA (Preferred Reporting Items for Systematic Reviews and Meta-analyses) guidance. But due to COVID -19, PROSPERO chose not to register any protocol of studies not focusing on the pandemic-related topics at the moment of registration.

The research team conducted a systematic search electronically, using the PubMed, Cochrane, and Embase databases, from October 2021 to September 2022. The search did not apply any restrictions to the year of publication. The participant’s age was limited to 18 years old. No limitations were applied to article types and language of publication. The search strategy in PubMed combined the subject headings and free-text keywords, (((("hospital s"[All Fields] OR "hospitalisation"[All Fields] OR "hospitalization"[MeSH Terms] OR "hospitalization"[All Fields] OR "hospitalising"[All Fields] OR "hospitality"[All Fields] OR "hospitalisations"[All Fields] OR "hospitalised"[All Fields] OR "hospitalizations"[All Fields] OR "hospitalized"[All Fields] OR "hospitalize"[All Fields] OR "hospitalizing"[All Fields] OR "hospitals"[MeSH Terms] OR "hospitals"[All Fields] OR "hospital"[All Fields]) AND ("patient s"[All Fields] OR "patients"[MeSH Terms] OR "patients"[All Fields] OR "patient"[All Fields] OR "patients s"[All Fields])) OR (("oncologic"[All Fields] OR "oncological"[All Fields] OR "oncologically"[All Fields] OR "oncologics"[All Fields]) AND ("patient s"[All Fields] OR "patients"[MeSH Terms] OR "patients"[All Fields] OR "patient"[All Fields] OR "patients s"[All Fields])) OR (("oncologic"[All Fields] OR "oncological"[All Fields] OR "oncologically"[All Fields] OR "oncologics"[All Fields]) AND ("disease"[MeSH Terms] OR "disease"[All Fields] OR "diseases"[All Fields] OR "disease s"[All Fields] OR "diseased"[All Fields])) OR ("genetic diseases, inborn"[MeSH Terms] OR ("genetic"[All Fields] AND "diseases"[All Fields] AND "inborn"[All Fields]) OR "inborn genetic diseases"[All Fields] OR ("genetic"[All Fields] AND "diseases"[All Fields]) OR "genetic diseases"[All Fields]) OR ("genetic diseases, inborn"[MeSH Terms] OR ("genetic"[All Fields] AND "diseases"[All Fields] AND "inborn"[All Fields]) OR "inborn genetic diseases"[All Fields] OR ("genetic"[All Fields] AND "disorders"[All Fields]) OR "genetic disorders"[All Fields]) OR ("chronic disease"[MeSH Terms] OR ("chronic"[All Fields] AND "disease"[All Fields]) OR "chronic disease"[All Fields] OR ("chronic"[All Fields] AND "diseases"[All Fields]) OR "chronic diseases"[All Fields]) OR (("patient s"[All Fields] OR "patients"[MeSH Terms] OR "patients"[All Fields] OR "patient"[All Fields] OR "patients s"[All Fields]) AND ("chronic disease"[MeSH Terms] OR ("chronic"[All Fields] AND "disease"[All Fields]) OR "chronic disease"[All Fields] OR ("chronic"[All Fields] AND "diseases"[All Fields]) OR "chronic diseases"[All Fields])) OR "oncology patient"[All Fields]) AND "Drug Therapy"[MeSH Terms] AND "Treatment Outcome"[MeSH Terms] AND ("drug resistance, bacterial"[MeSH Terms] OR (("anti infective agents"[Pharmacological Action] OR "anti infective agents"[MeSH Terms] OR ("anti infective"[All Fields] AND "agents"[All Fields]) OR "anti infective agents"[All Fields] OR "antimicrobial"[All Fields] OR "antimicrobials"[All Fields] OR "antimicrobially"[All Fields]) AND ("resist"[All Fields] OR "resistance"[All Fields] OR "resistances"[All Fields] OR "resistant"[All Fields] OR "resistants"[All Fields] OR "resisted"[All Fields] OR "resistence"[All Fields] OR "resistences"[All Fields] OR "resistent"[All Fields] OR "resistibility"[All Fields] OR "resisting"[All Fields] OR "resistive"[All Fields] OR "resistively"[All Fields] OR "resistivities"[All Fields] OR "resistivity"[All Fields] OR "resists"[All Fields])))). The following string was used in Embase, (’treatment outcome’/exp OR ’treatment outcome’) AND (’antibiotic resistance’/exp OR ’antibiotic resistance’) AND (’hospital patient’/exp OR ’hospital patient’ OR ’oncology patient’ OR ((’oncology’/exp OR oncology) AND (’patient’/exp OR patient)) OR ’chronic disease’/exp OR ’chronic disease’ OR ’genetic disorder’/exp OR ’genetic disorder’ OR ’oncological disease’ OR (oncological AND (’disease’/exp OR disease)) OR ’patients chronic diseases’ OR ((’patients’/exp OR patients) AND chronic AND (’diseases’/exp OR diseases))) AND (’drug therapy’/exp OR ’drug therapy’) AND ([adolescent]/lim OR [child]/lim OR [infant]/lim OR [newborn]/lim). Before further search with a broader string #17 AND ’article’/it AND ([adolescent]/lim OR [child]/lim OR [fetus]/lim OR [infant]/lim OR [newborn]/lim OR [preschool]/lim OR [school]/lim) AND (’adverse device effect’/lnk OR ’adverse drug reaction’/lnk OR ’disease management’/lnk OR ’drug combination’/lnk OR ’drug concentration’/lnk OR ’drug interaction’/lnk OR ’drug resistance’/lnk OR ’drug toxicity’/lnk OR ’side effect’/lnk OR ’special situation for pharmacovigilance’/lnk OR ’therapy’/lnk OR ’unexpected outcome of drug treatment’/lnk) and in Cochrane ’antibiotic resistance’/exp/mj AND ([cochrane review]/lim OR [systematic review]/lim OR [meta analysis]/lim OR [randomized controlled trial]/lim OR ’controlled clinical trial’/de) AND [article in press]/lim AND ([english]/lim OR [french]/lim OR [italian]/lim) AND ([embryo]/lim OR [fetus]/lim OR [newborn]/lim OR [infant]/lim OR [child]/lim OR [preschool]/lim OR [school]/lim OR [adolescent]/lim) AND [humans]/lim AND [abstracts]/lim AND [clinical study]/lim AND ([embase]/lim OR [medline]/lim OR [pubmed-not-medline]/lim) AND [medline]/lim was the label used. Extended literature hunting in the CMI database: antimicrobial resistance children OR impact OR on OR therapy OR surveillance OR control OR management OR stewardship "immune depressed" was also made.

The modified PECO inclusion/exclusion criteria (Table 1) served to screen titles/abstracts and complete texts.

**Table 1:** Inclusion/exclusion criteria applied

| Criteria | Inclusion | Exclusion |
| --- | --- | --- |
| Population | Humans<br>All sexes<br>Aged 0 to 18 years<br>Infections with AMR organisms and ESBL producing organisms.<br>Post-antibiotic era studies on infected populations | Plants only<br>Animals only |
| Outcomes | Associated with immune-depression (relative state of fragile health status requiring constant or ongoing care).<br>Impact on ongoing therapy at time of infection | Health-related quality of life only<br>Outcomes associated with the evaluation of the intervention only<br>Reports on analytic methodologies |
| Study design | Case-control studies<br>Cohort studies<br>Cross-sectional studies<br>Longitudinal studies<br>Randomised control trials<br>Modelling studies<br>Economic evaluations<br>Evaluations of intervention reporting impact of exposure of fragile children to AMR | Letters<br>Reviews<br>Editorials<br>Conference reports<br>Case series reports<br>Evaluations of interventions not reporting impact of exposure |
| Language | English<br>French<br>Italian | All other unfamiliar language to the reviewer |

### 2.4. Data selection and collection process

All reviewers screened the references (Titles and Abstracts) individually, and disagreements were resolved through discussion with mates investigating in the same field and consensus with co-authors. The primary reviewer used prevalently manual screening tools like Excel to select the qualifying articles. Like in previous systematic reviews, titles and abstracts of the retrieved documents were initially assessed, and other documents non-related to the topic were excluded. For data from the grey literature, executive summaries, tables of contents, and documents (whichever was available) were screened. The full text of potentially eligible documents was then obtained and assessed for relevance or duplication against predefined selection criteria. Searches were limited to English, Italian and French articles to avoid unreliable translations. Duplicates were removed with Zotero.

Selected literature was screened for relevancy to the population (humans), antibiotic resistance of investigation, pathogens involved in the study, type of resistance mechanism, and information on mortality and LoS. Reviewers strive to understand whether it was an excellent idea to combine the treatment or not (where necessary), whether the use of a different antibiotic resulted in being in favor of the patient/population, whether following the primary patient’s / population therapy, ignoring the resistant bacterial infection was the preferred choice, and why?

### 2.5. Data collection process

Two (D. D. and I.N.W.) reviewers independently examined the selected articles at the primary stage of the process. The PRISMA RoB 2 and ROBINS tools were used to assess the risks of bias in the selected literature.

Data collected included studies focusing on children, the average time of stay in hospital of the patients, the average mortality and morbidity rate, and the impact of the resistant bacterial infection on the outcome of the patient, in association with the type of bacterial resistance, immunodepression and the AMR of question.

Data on mortality rate was retrieved from the studies as much as possible, as a significant outcome of the burden of AMR, together with information on the Hospital’s length of stay (LoS) upon detection of the resistance; LoS is a significant contributing factor to hospital increased costs (45). It is prudent not to underestimate the cost of hospitals’ consumables, as they are always part of a budget.

We did not request ethical approval for this work since the data analyzed are not directly related to individuals.

### 2.6. Data analysis

We screened the selected studies according to the following criteria: study type, populations, pathogens, AMR, immunodepression, and reporting of the mortality and LoS in the hospital during therapy. The analysis was based on comparing the percentage of different categories and subcategories under revision to understand what exactly is going on in the pediatric world of immune-depressed children contracting resistant infections.

## 3. Results

About 350 unique titles and abstracts were retrieved from October 2021 to September 2022. Applying the selection process resulted in 110 studies being included in the final review, as appears on the PRISMA workflow diagram (Figure 1). The articles were then assessed for bias according to the PRISMA guidelines for the writing-up of systematic review style using the RoB II analytic tool for all randomized controlled trials included in the review. The risk of bias assessment generated an overall positive analysis resulted in being low (Figure: 1a). Most of the induced bias arose from the scarcity of information on pediatric populations, mortality, and LoS in hospitals during events of AMR infections, as it appears on both the bar chart and the traffic light plot (Figure: 1b).

**Figure 1:**
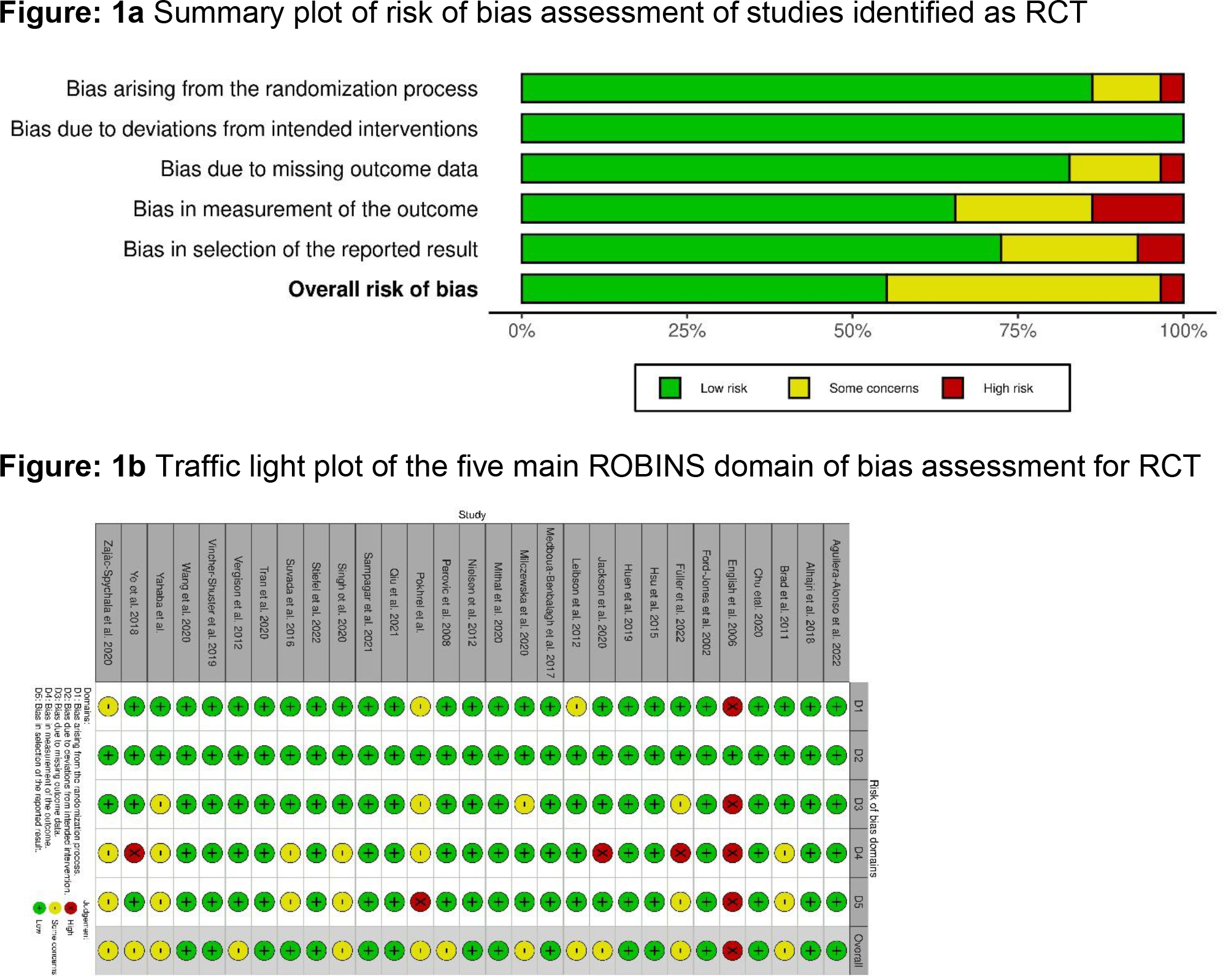
PRISMA flow diagram See pdf

**Figure 2:**
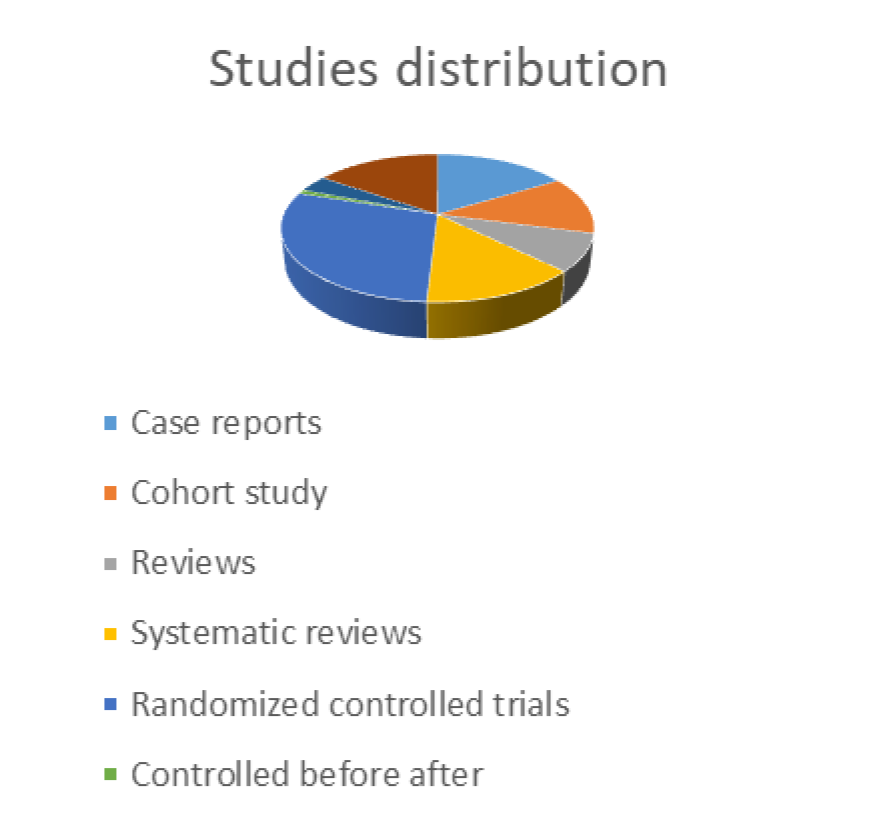

The articles were pooled for information relative to the current review scope: population, pathogens of concern in the study, type of resistance mechanisms of matter, immune kind of depression represented among the people, and the mortality and morbidity rate eventually reported. Retrieved information was classified into tables and described in figures (Pies distribution charts) in Microsoft Excel 2019.

From the 110 studies selected for the review, the more numerous types of studies, with 29 % of the total (Table 2), were randomized controlled trials. Although it was successfully possible to analyze the articles from the types, some (17, thus 15%) could not be accurately identified.

**Table 2:**
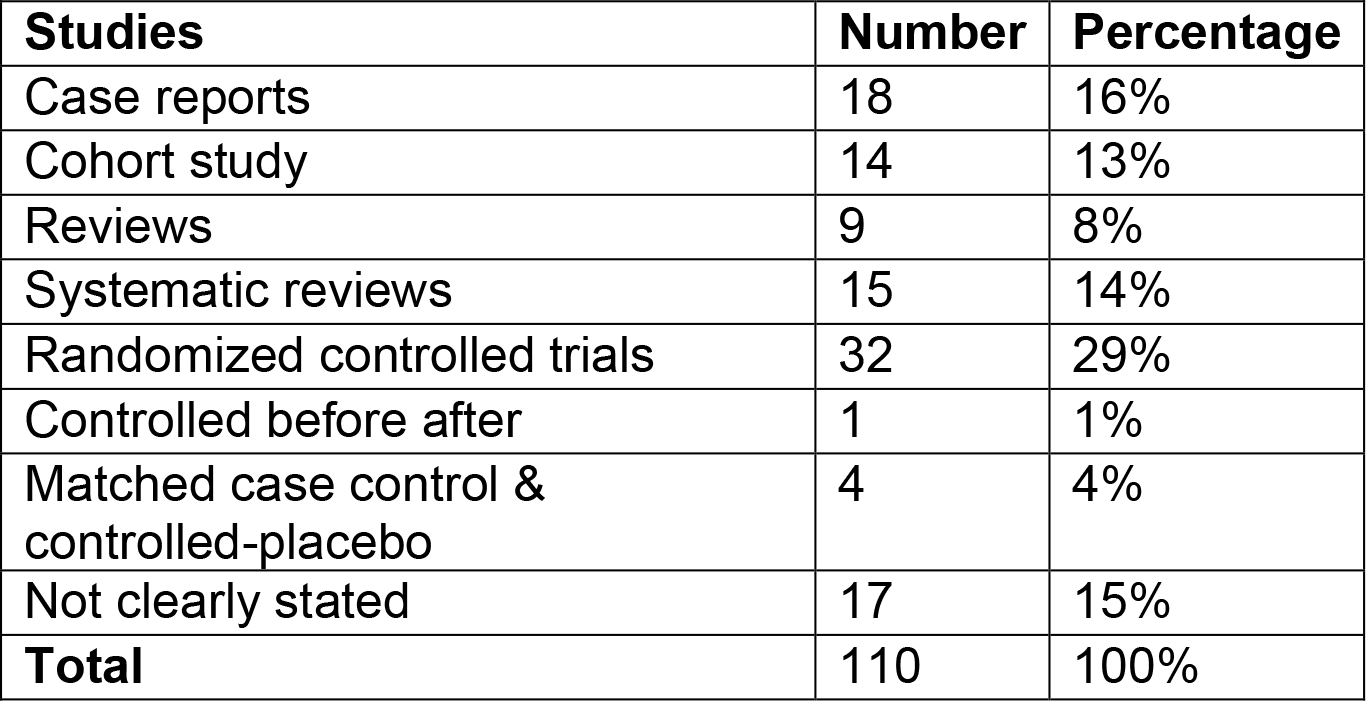
Estimate number of studies per categories

From the following table though (Table 3: Population type), it was evident that only 45, thus 41% of the same articles were studies on pediatric populations; therefore, studies that retrieved information were more specific to the present review than the others—searching for this information among the selected studies led to the discovery that about 50% of them, around 55 (Table 3), needed to have the study population type clearly defined, as represented in Figure 3. This can be explained by the fact that most studies did instead report the pathogens of concern (Table 4) and the resistance of investigation in the scope of the study (Table 5). In fact, from the 45 pediatric studies, only 8, thus 18 % of them, did not state which type of pathogen was of concern in their studies (Figure 4), versus a relatively low number of 10 or 22%, on the other hand, which did not mention the type of resistance mechanisms (Figure 5). From these two figures, it is visible that current studies report gram–negative pathogens as the most commonly reported or encountered among children compared to gram-positive organisms and others (mycoplasma, fungi, etc…), followed by the infections caused by mixed pathogens. MDR is the most common type of resistance reported in studies (Figure 5), which seems to be explained by the high number of infections caused by both gram-negative bacteria mainly and mixed pathogens (Table 4). Broad-spectrum antibiotics are often used in these situations, which may become favorable for resistances, and why not, of multiple types. The current figure (Figure 5) shows how the reporting of AMR among pediatrics is distributed so far in the world. Few studies report single AMR (2%) to antibiotics like colistin, ampicillin, fluoroquinolones, penicillin, third- generation cephalosporins, and other rare phenomenon–related - resistance types, like chemoresistance or drug-associated diarrhea. But suppose the among of MRSA seems to be low. In that case, this might be because some other MRSA (7% alone in this case), when reported together with some other resistance mechanisms revelated in the same study, ended up being counted under the MDR identified per study. The number of ESBL found in this population (4%) is not insignificant either, for it is known that these types of infections are very lethal in children. The estimative distribution of the data mentioned above (Figure 5) is enough of a red flag to set the alarm on the necessity of better controlling the situation at a global level.

**Figure 3:**
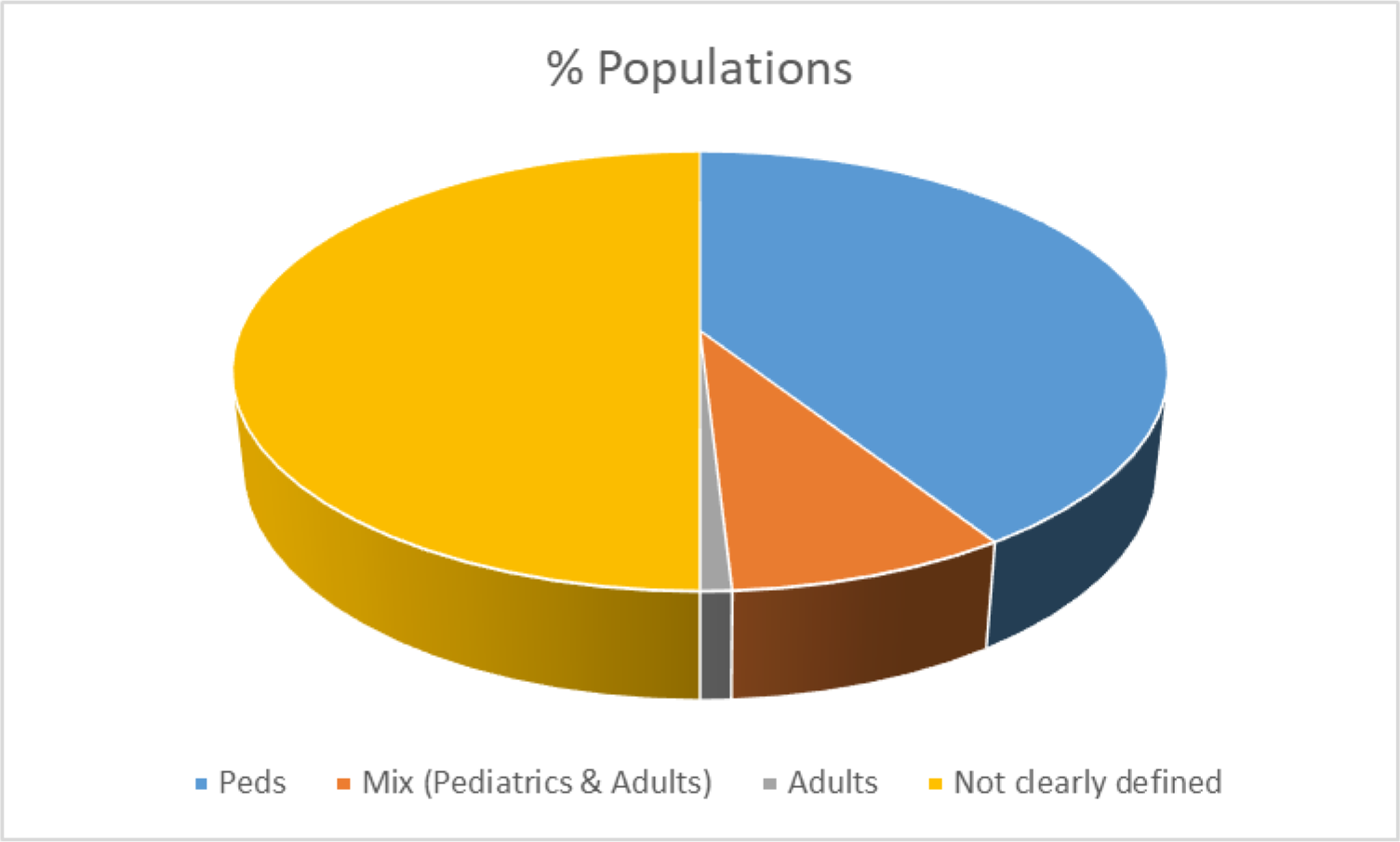

**Figure 4:**
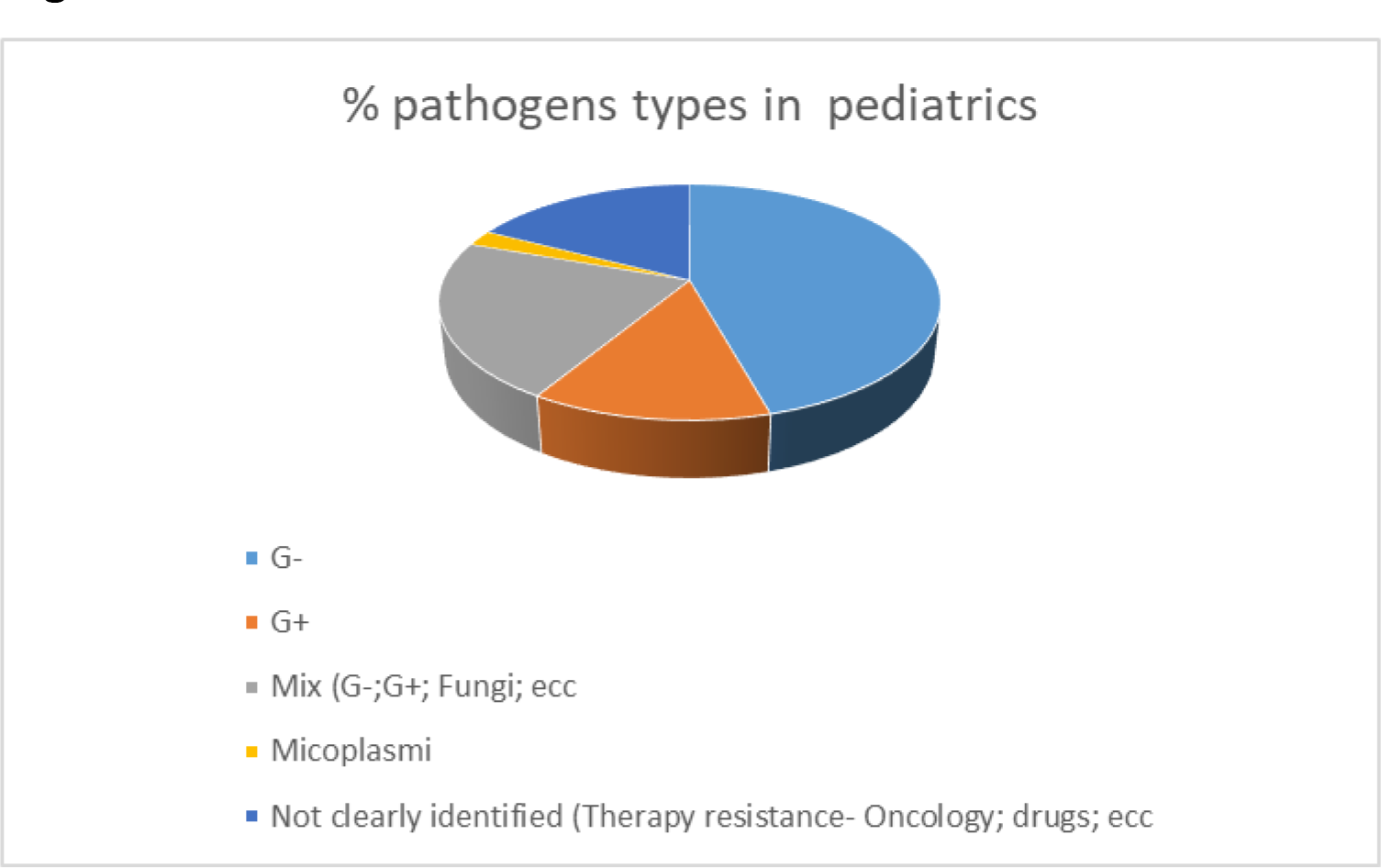

**Figure 5:**
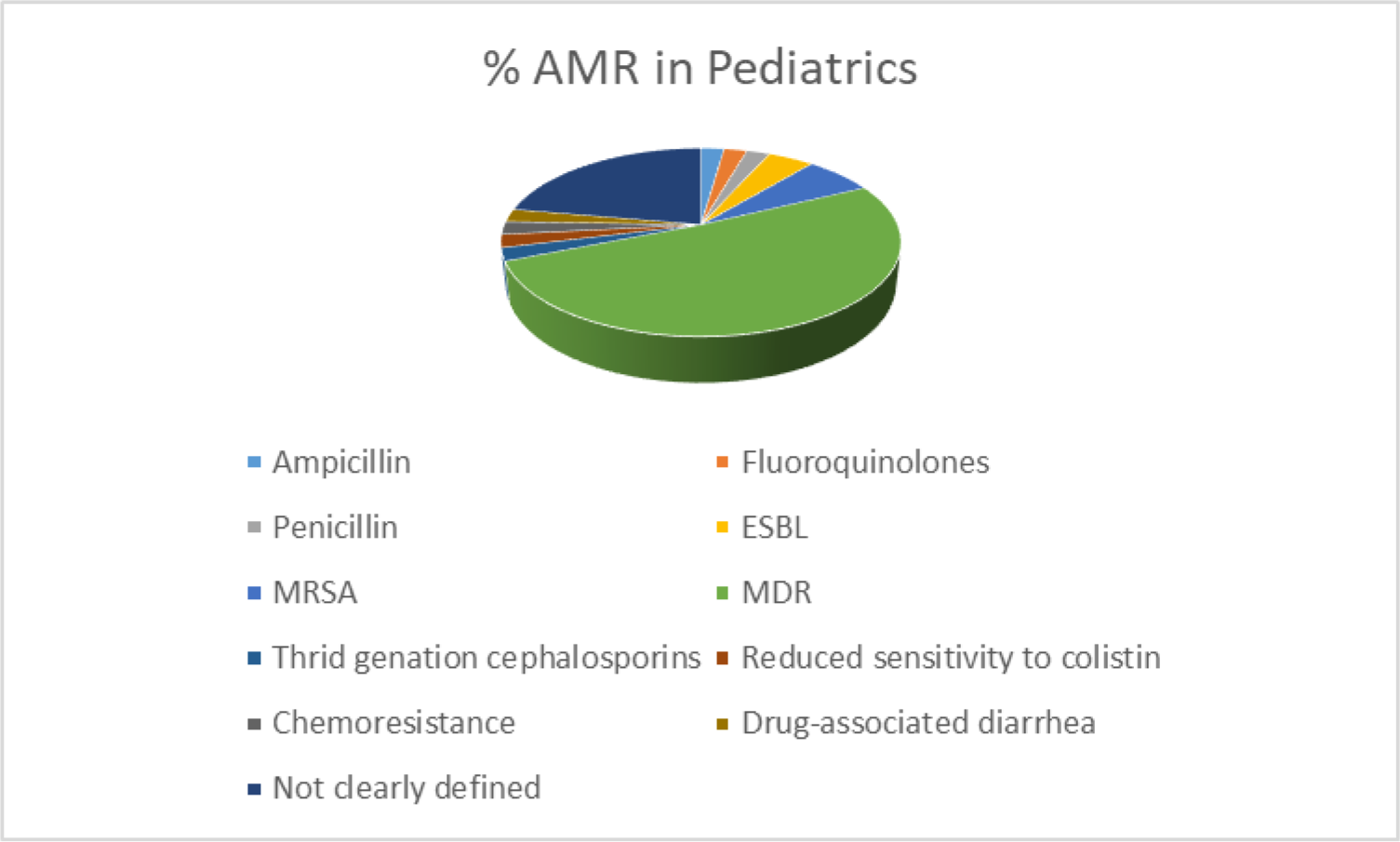

**Table 3:** Estimate population of study type, per studies

| Populations | Number | Percentage |
| --- | --- | --- |
| Peds | 45 | 41% |
| Mix (Pediatrics & Adults) | 9 | 8% |
| Adults | 1 | 0,01% |
| Not clearly defined | 55 | 50% |
| **Total** | **110** | **100%** |

**Table 4:** Estimate repartition of resistant bacteria per categories

| Pathogens | N# | Percentage |
| --- | --- | --- |
| G- | 21 | 47% |
| G+ | 6 | 13% |
| Mix (G-; G+; Fungi; ecc | 10 | 22% |
| Micoplasmi | 1 | 2% |
| Not clearly identified (Therapy resistance- Oncology; drugs; ecc | 8 | 18% |
| Total | 45 | 100% |

**Table 5:** Estimate repartition of resistance per studies screened

| Resistance | N# | Percentage |
| --- | --- | --- |
| Ampicillin | 1 | 2% |
| Fluoroquinolones | 1 | 2% |
| Penicillin | 1 | 2% |
| ESBL | 2 | 4% |
| MRSA | 3 | 7% |
| MDR | 23 | 51% |
| Thrid genation cephalosporins | 1 | 2% |
| Reduced sensitivity to colistin | 1 | 2% |
| Chemoresistance | 1 | 2% |
| Drug-associated diarrhea | 1 | 2% |
| Not clearly defined | 10 | 22% |
| **Total** | **45** | **100%** |

Further investigations on the types of immune depression encountered among the reported patients’ populations describes a high number of genetic diseases and acquired severe pathologies as the leading cause of immune suppression in children. Among the studies focusing on pediatric retrieve during the study period, only one reported on a case of immune depression associated with organ transplantation or hospitalization in ICU / Trauma units; 16% of the studies were on neonates, generally considered as fragile from birth till the sixth month of growth, and what the present study chose to call “hospitalized children” (Table 6), referring to those having six months onwards. It is also possible the note that even in this case, 20% of the studies couldn’t clearly state the type of disease associated with the specific study population (Figure 6).

**Figure 6:**
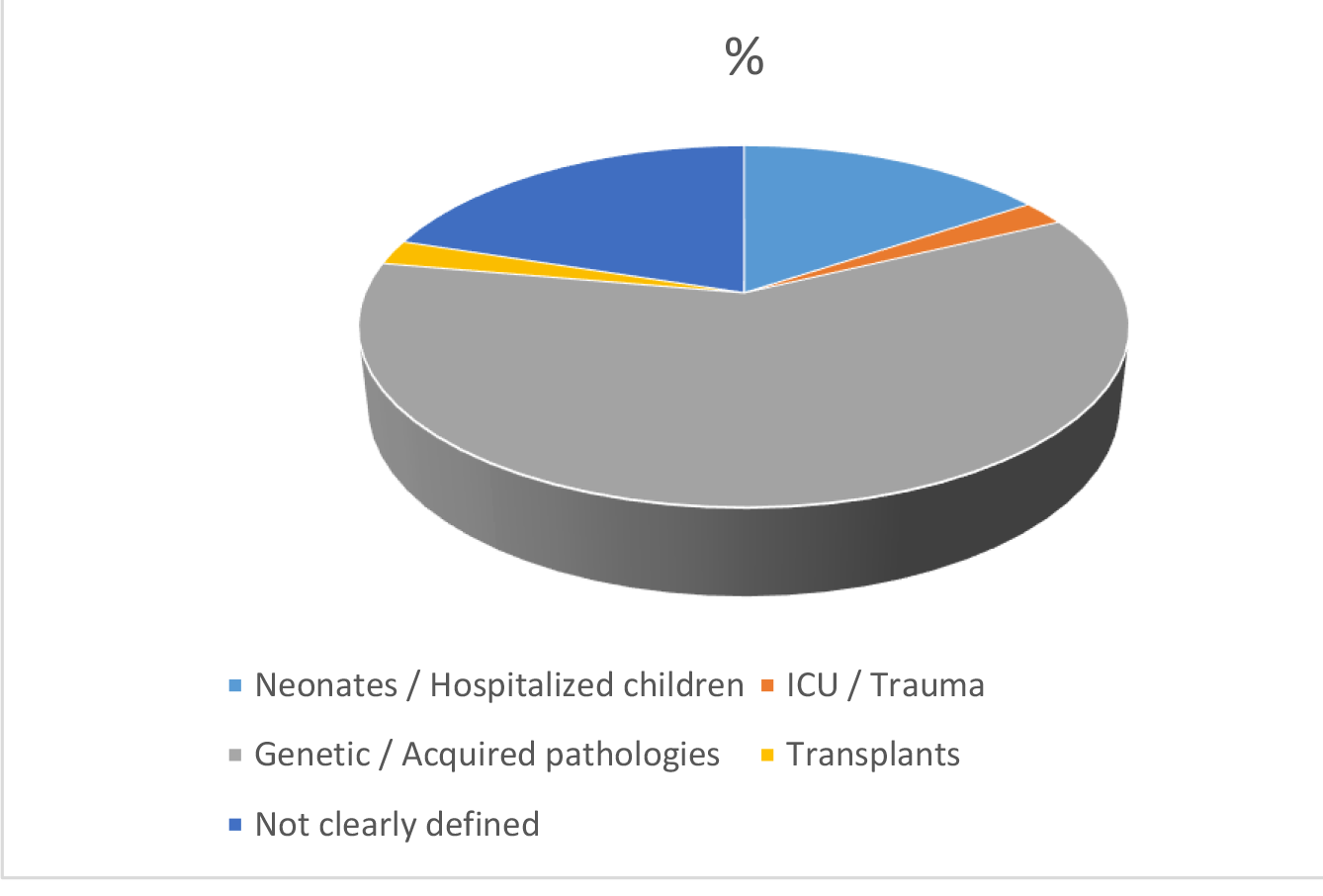

**Table 6:** Estimate repartition of immune-depression types in studies screened

| Immunodepression | N# | Percentage |
| --- | --- | --- |
| Neonates / Hospitalized children | 7 | 16% |
| ICU / Trauma | 1 | 2% |
| Genetic / Acquired pathologies | 27 | 60% |
| Transplants | 1 | 2% |
| Not clearly defined | 9 | 20% |
| **Total** | **45** | **100%** |

An attempt to evaluate the burden of AMR on the mortality and length of stay in hospital in the pediatric population from the studies mentioned above yielded the discovery that most research in the pediatric camp does not aim at investigating the duration in a hospital or the mortality rate among children, due to AMR. About 56% (Table 7) of them do not mention this information at all in the study; about 36% do declare a decrease in death and duration of hospitalization after infections caused by AMR bacteria associated with AMS programs and other surveillance strategies against AMR infections; 2 of 45 studies do report a noted increased mortality rate in the cause of the study due to the infections; and about the same number of studies reported on the current o as a deviated outcome, either as a subsequent one (Figure 7).

**Figure 7.**
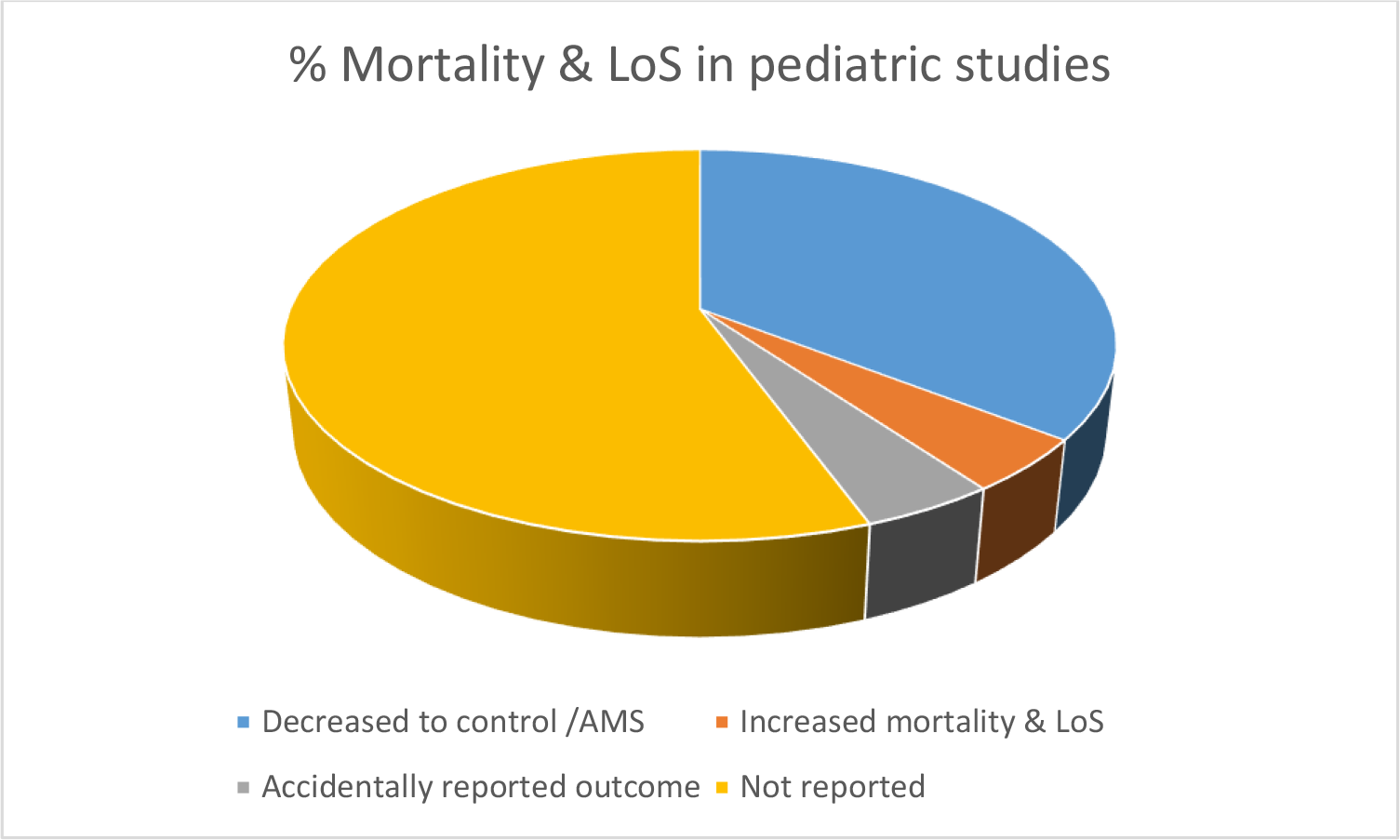

**Table 7.**
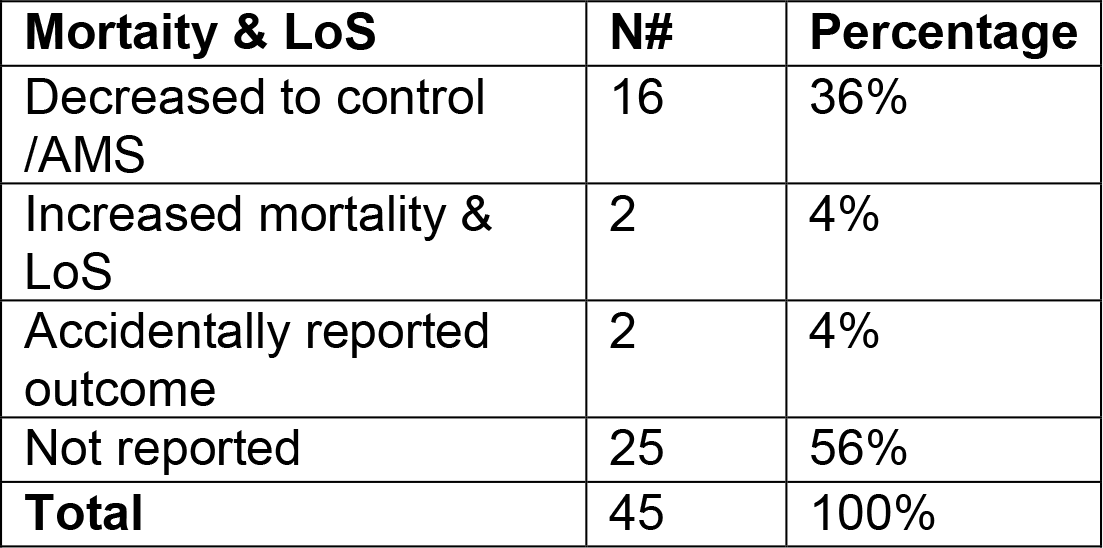
Estimate repartition of mortality and Length of hospital stay (LoS) in studies

## 4. Discussion

The present systematic review of studies aimed to search and present what in the peer- reviewed literature is the current knowledge of the impact resistant bacterial infections have on children in general, but on fragile kids, in particular. Then, based on the statistical results found, estimate the necessity to repropose and undertake practical actions suggested by experts, like doing more research on the topic (7,75) and providing as much data as possible at various levels (76). This is mainly done through observational studies (prospective and retrospective), randomized controlled trials, and case controls to raise public health concerns, participation, and exemplary implementation of antimicrobial stewardship programs (AMS) where necessary.

It was found that studies address the burden of AMR, mainly when a particular case is found. The issue reported will further generate a specific interest in the particularities of the experience, like the patient category (sex, age, nationality, employment type, health status) and hospital/community of concern. Therefore, some trials followed up, together with some cohort and case-control studies.

Results presented in this review describe that out of about 350 studies, only a few focused on the impact of antimicrobial resistance in pediatrics (45; Table 3). However, the quality of the studies was overall good, from the bar chart of risk of bias assessment of studies identified as RCT and other assessment tools used for further studies.

The variety of isolates of concern among children (Table 4) may indicate how urgent it is to protect fragile pediatric patients yet from most pathogens, but more especially from gram- negative bacteria already known to be able to develop resistance against antibiotics. The pseudo-list of antibiotics observed in Table 5 indicates how many of these drugs are in need also among kids. When is already being emphasized that they are of few benefits to their health. But the necessity of using them is visible, especially the amount of MDR infections.

This study showed that most infected patients in this category are fragile children affected by genetic conditions or certain severe diseases acquired during their life. This might explain why they are so easily threatened by gram-negative pathogens (77,78). These are often encountered in hospital standard wards and fewer in intensive care units, like trauma or ICUs, which are, in general, much more sterilized units (22,46).

Precise data on the mortality and morbidity rate (79) of fragile pediatric patients after hospitalization is almost impossible, if not wholly absent from previous work. A last review on a similar topic affirms that researchers agree that AMR increases mortality and length of hospital stay in about 50 % of the studies(79–85). Researchers seem to be convinced that there is an evident increase in mortality among this patient group (hospitalized children). Still, precise data are necessary to evaluate to which extent. To avoid inappropriate prescribing of antibiotics or administration, specific centers carefully adhere to collaboration programs for the control, management, and surveillance of antibiotics, which cultivate a cautious use of them, and therefore lessen the insurgence of resistance(61,63,64,73,75,86). Several of these centers report successful implementation of ASP in their facilities. When possible, they also publish work on successful therapies against resistant bacteria, which is very important for health care and public knowledge of the status of AMR in general(87–97). The latter may explain why about 36% (Table 7) of the studies screened reported a decrease in mortality and LoS in hospitalized immune-depressed kids during their experimentation.

## 5. Conclusion

This study confirms the need of relevant information on the impact of AMR in hospitalized immune-depressed children. This will be helpful in the understanding of how to control the emergency of resistances in and out of hospital settings. These will also be very helpful in knowing how to handle the therapy of these patients, such as not to create more harm than to cure them, and to control the cost associated with the treatments involved in the cure of these patients, with the extensive use of antibiotics in general and in particular, where possible and where necessary. Hospitals and laboratories engaged in stewardship programs should collaborate in continuation to share and promote successful therapies and experiences as there seem to be ways to bypass antimicrobial resistance keeping an eye on the dosages and type of combination therapy to implement (98). Further review may be run to perform a meta-analysis of the estimated burden of AMR infections in pediatrics to confirm the theory in the literature. Sensibilization campaigns for vaccinations need to be promoted accordingly, whenever necessary, but more importantly, taught and prepared well in advance by well-trained local and international professionals to avoid consequential limitations to the programs(1,58,58,99–105). Relevant suggestions for managing community immunocompromised patients should be considered and implemented where and when possible (106,107).

## Supporting information

PRISMA CHECKLIST

Pediatric study reports screened

## Data Availability

All data produced in the present study are available upon reasonable request to the authors.

---

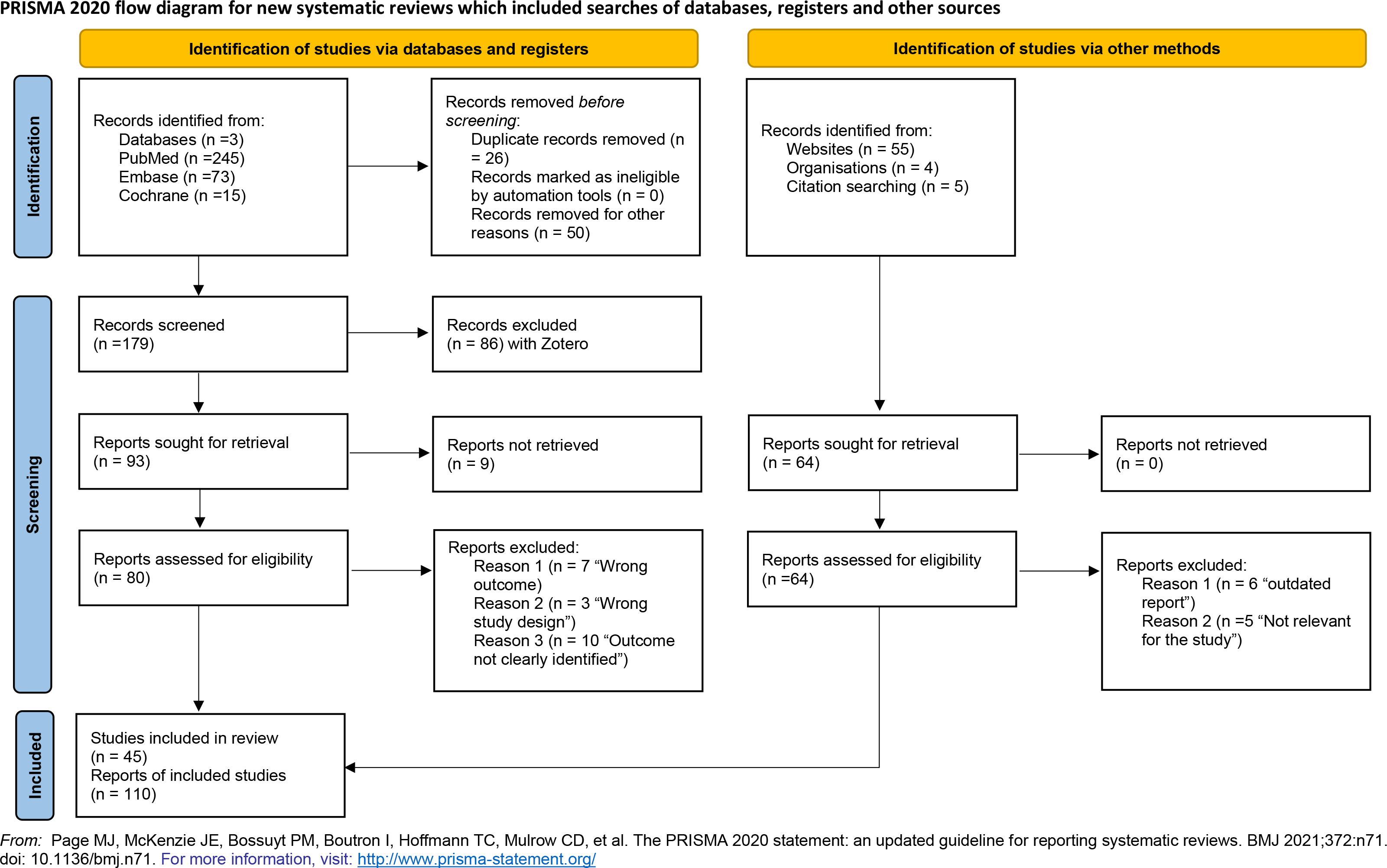

Table 8: Data synthesis

See pdf

