## Supplementary material for "Surveillance of the Impact of Antimicrobial Resistant infections in Immunosuppressed Children’s therapy: A systematic review": Pediatric study reports screened

| Author and year | Study design | Study Location | SP | Type of resistance | Pathogens | Type of antibiotics | Type of immunodepression | Mortality & LoS |
|---|---|---|---|---|---|---|---|---|
| (1) | Case report | France | Peds | MDR | Pandoraea sp. | | | |
| (2) | Case report | Malaysia | Peds | MDR | Burkholderia pseudomallei G- | | Cystic fibrosis | D |
| (3) | Case report | South Africa, Johannesburg | Peds | MDR | Psedomonas spp. | | | |
| (4) | Case report | South Africa, Pretoria | Peds | | | | | |
| (5) | Case report | UK | Peds | MDR | Serratia marescens & S. aureus | | X-linked chronic granulomatous disease (CGD) | D |
| (6) | Case report | USA, Canada, and Australia | Peds | None - Reduced sensitivity | B. pseudomallei | agressive treatment regimen | Cystic fibrosis | D |
| (7) | Case report | | Peds | MDR | P. aeruginosa | | Severe aplastic anemia | |
| (8) | Case report | | Peds | Fluoroquinolone | U. urealyticum | Azythromycin & doxycycline | Kidney transplant | |
| (9) | Case report | | Peds | MDR | | | Acute otitis media | D |
| (10) | Case report | | Peds | MDR | Pandoraea | | Cystic fibrosis | |
| (11) | Case report | | Peds | No resistance | Staph. Pseudintermedius | Methicillin | Anaplastic ependymoma | |
| (12) | Cohort study | | Peds | P. aeruginosa; ESBL producers | Gram -negative | Empiric antibiotic therapy | Neaonates intensive care unit | D |
| (13) | Cohort study | | Peds | MDR including ESB | Enterobacter cloacae | | ICU | |
| (14) | Cross-sectional study | Nepal | Peds | First-line antibiotics like ampicilin | For pneumonia | Ampicillin + ceftriaxone + others | N/HC | D |
| (15) | Propective osservational cohort study | European | Peds | MRSA | Gram-positive & gram-negative (-50%; Klebsiella & Acinetobacter spp.) Candida spp. 7% | | | |

| | | | | | | | | |
|---|---|---|---|---|---|---|---|---|
| (16) | Prospective , multi-site, cohort study | Canada | Peds | | H. influenzae & M. catarrhalis, S. pneumoniae | | Acquired infection | |
| (17) | Case report | USA | Peds | MRSA | | Vancomycin | Cystic fibrosis | D |
| (18) | Prospective multicenter study | Uganda | Peds | MDR | G- & G+ | Multiple | Oncologic patients | I |
| (19) | Prospective observational study | | Peds | | Pseudomonas spp. | | Cystic fibrosis | |
| (20) | Prospective study | | Peds | Third generation cephalosporins | >G-; Extended b-lactamases producers | Cefoperazone-sulbactam or carbapenems | Liver disease-related ascites | I |
| (21) | RCT | India | Peds | GPC and E. coli, Kliebsiella spp.; P. aeruginosa | Mix pathogen | | Oncologic patients | D |
| (22) | RCT | China | Peds | Multiple | K. Pneumoniae | | Neonates | D |
| (23) | RCT | | Peds | MDR | Gram-negative | | Neurogenic bladder | D |
| (24) | RCT | | Peds | Antibiotic associated diarrhea | Pseudomonas spp. | cephalosporin s | Pseudomonas associated diarrhea | |
| (25) | RCT | Spain | Peds | Methicillin | S. pyogenes & S. pneumoniae | | Hospitalized children | |
| (26) | RCT | | Peds | MDR | Gram-negative | | Urinary tract infection | |
| (27) | RCT prospective | Taiwan | Peds | MDR VAP | MDR-VAP | | VAP infection & | D |
| (28) | RCT, prospective | Europe | Peds | MRSA | Gram-positive & gram-negative (-50%; Klebsiella & Acinetobacter spp.) Candida spp. 7% | | Cystic fibrosis; toxic shock syndrome; skin and soft tissue infections | |
| Liebson et al. 2012(29) | RCT, prospective | Israel | Peds | Piperaciclin & Amikacin | CONS bacteremia; Acinetobacter & K. Spp; Candida spp. | | Oncologic patients | D |
| (30) | Retrospective cohort study | Japan | Peds | MDR | For Community acquired pneumonia (CAP) | B-lactams; tetracycline and quinolone | None | D |
| (31) | Retrospective study RCT | | Peds | MDR | | | Hematological malignancies | |

| | | | | | | | | |
|---|---|---|---|---|---|---|---|---|
| (32) | Review | Madrid, Spain | Peds | MDR | Non salmonella typhi | | 11 underlying immunedepression: malignancy; IgA-IgG2 deficit; chronic granulomatous disease; HIV; Systemic lupus erythematosus; liver disease; hypoxic hypoxic - ischemic encephalopathy | |
| (33) | Systematic review | | Peds | MDR | | | Cystic fibrosis | AO |
| (34) | | | Peds | | P. aeruginosa | S: Colistin | N/HC | AO |
| (35) | | Benin | Peds | ESBL-E | K. pneumoniae; E. coli | | Oncologic patients | |
| (36) | | | Peds | | | | Malignancies and haematopoietic steem cell transplant | |
| (37) | | | Peds | MDR | CA-MRSA | Clindamysin Vancomycin | | |
| (38) | | Ghana | Peds | MDR | Non typhoidal salmonellae ; S. aureus; S. pneumoniae; Salmonella ser. Typhi | | | |
| (39) | | | Peds | MDR | Mix pathogen | | Haematopoeitic stem cell transplantation | D |
| (40) | | | Peds | Penicillin-resistant | Aerococcus viridans | | Sickle-cell anemia | |
| (41) | | Taiwan | Peds | | | | Congenital anomalies | |
| (42) | | | Peds | MDR-A. baumannii | Gram-negative | Tigecycline | | |
| (43) | | | Peds | | | | Oncologic patients | D |
| (44) | | | Peds | MDR | Mix pathogen | | Children | D |
| (45) | | | Peds | | Viridans group streptococci (VGS) | | Immunocompromised | |

13. Alhajri N, Hamdy R. The epidemiology and outcomes of enterobacter cloacae bloodstream infections in children. Open Forum Infect Dis. 2018;5((Alhajri N.) Epidemiology and Biostatistics, George Washington University, Washington, DC, United States):S726.

26. Mithal LB, Otero S, Sun S, Arshad M. Trends in antibiotic resistance among uropathogens in the pediatric population: A single center experience in the us. Open Forum Infect Dis. 2020;7(SUPPL 1):S695.
